# Drivers and barriers to promoting self-care in individuals living with multimorbidity: a cross-sectional online survey of health and care professionals

**DOI:** 10.1101/2023.12.21.23300404

**Authors:** Susan Barber, Benedict Hayhoe, Sonia Richardson, John Norton, Manisha Karki, Austen El-Osta

**Author notes:** Corresponding author Susan Barber.

## Abstract

**Objective:** Investigate knowledge, attitudes, and perceptions of health and care professionals (H&CPs) in England concerning drivers and barriers for promoting self-care in service-users with multimorbidity.

**Design:** A cross-sectional online survey of the health and care workforce.

**Setting:** Health and social care workforce.

**Participants:** Eighty-eight health and social care professionals in England.

**Methods:** A cross-sectional online survey administered via Imperial College Qualtrics platform. Questions were asked about perceived drivers and barriers to promoting self-care in individuals with multimorbidity, including mental health.

**Results:** Extant barriers associated with service-users ability and opportunity to self-care were feelings of loneliness and social isolation (18.9%; n=61), mobility and access issues (14%; n=45). Strategies deployed by H&CPs to support self-care were social prescribing (17.9%; n=59), helping service-users to monitor their symptoms (15.2%; n=50), referring to recognised programmes to support self-management (13.9%; n=46), knowledge and understanding about the benefits of self-care (91.8%; n=67), the purposes of prescribed medicines (83.3%; n=60), and support for self-care (91.7%; n=66) were considered key drivers of successful self-management and to engagement between service-users and service providers. Service providers’ reported gaps in their knowledge including how to improve practical interactions to sustain health seeking behaviours by service-users (30.6%; n=49), health coaching (21.9%; n=35), improved understanding about effective self-care interventions (21.3%; n=34) and improving self-care in relation to medicines use (20%; n=32). Most respondents (92.2%; n=71) reported that the COVID-19 pandemic had highlighted the need for self-care, and (42.7%; n=32) agreed that the pandemic had a positive impact on their ability to promote self-care among service-users.

**Conclusions:** Self-care is important for service-users who live with multimorbidity. H&CPs are in a unique position to influence lifestyle choices and health-seeking self-care behaviours. Raising awareness about the importance of self-care, health literacy, knowledge, understanding and skills among service-users and providers is key to improving supported self-care.

## Introduction

Increasing numbers of people live with multimorbidity (MM), defined as the coexistence of two or more chronic health conditions in the same individual (1–3). Service-users with multimorbidity must cope with a complex array of associated issues, including the effects of the health conditions themselves, a higher treatment burden, polypharmacy, multiple health care appointments, psychosocial issues, and stress (4,5).

Self-care may assist individuals with multimorbidity in meeting some of these needs. Among the positive outcomes of supported self-care for individuals living with MM are improved self-efficacy, independence in daily activities, quality of life, goal achievement (6), improved depression scores (7), diet (8), and composite measures of glycaemic control, blood pressure, lipids, and depression scores (7,9). Of particular importance in the context of stretched health services is the potential for reduced use of health services, hospital admissions and emergency services (10–12), and prevention of illness progression (13).

Providers of health and social care utilise a variety of interventions to support individuals with multimorbidity, including the deployment of strength-based approaches (14–16), the promotion of self-care, and self-management strategies (17). Examples include access to local assets such as local community services, information and support, signposting and social prescribing, use of shared decision-making, and alternative person-centred approaches to promote patient activation and self-care (14–17).

Health and care professionals (H&CPs) who are well trained and organised are likely to respond better to the challenges of supporting people with multimorbidity to self-care (13). There is a need to reform the way the healthcare system can deliver care, train, and re-skill H&CPs, with a focus on the development of necessary working relationships designed to support service-users to self-care (18,19). Multiple barriers have been identified to supporting self-care among individuals with multimorbidity (20–23), such as complex psychosocial and cultural barriers and practicalities relating to roles and responsibilities among service-users and health and social care providers (24).

### Study rationale

There is little comparative research available to guide our understanding of what the health and social care workforce consider as drivers and barriers to good quality supported self-care for people who live with multimorbidity. Understanding how the self-care agenda is being supported by H&CPs is important. Whilst many studies have focused on challenges and outcomes of self-care interventions (6–11,13), to our knowledge this is the first to focus on the knowledge, attitudes and perceptions of H&CPs about extant barriers. National guidance calls for H&CPs to support self-care as part of personalised care across the health and care system (18,25), whilst international guidance calls for personalised and supported self-care to be considered as a key to improving patient safety (2). National guidance suggests that this requires a shift in relationships between H&CPs and people, in which all the person’s health conditions and needs are considered together, and in which that person’s own skills, strengths and attributes are taken into account (18). Little is known about the additional needs of H&CPs to help deliver this.

### Study objectives

This study aimed to investigate the knowledge, attitudes, and perceptions (KAP) of H&CPs in England concerning extant barriers and drivers to promote self-care among service-users with multimorbidity.

## Methods

### Study design

We conducted a cross-sectional online survey of H&CPs in England about their knowledge, attitudes, and perceptions (KAP), drivers and barriers to promoting self-care in service-users.

The link to the electronic open survey was published and available on the Imperial College Qualtrics platform between 5 February 2021 and 18 August 2021; the survey could be accessed by anyone with a link. Potentially eligible participants received an invitation email, including a Participant Information Sheet and link to the survey, from the study team; the first question in the survey requested informed consent from the participant after which the survey could be completed. Completion of the survey was voluntary. No incentives to complete it were offered. Co-investigators also disseminated the email to their professional networks. The data collected were stored on the Imperial College London secure database, and only the team researchers could access the survey results.

The survey comprised a total of 17 questions and was accessible using a personal computer or smartphone. The questions were displayed on one page with a progress bar. Some questions invited respondents to tick more than one response where that was appropriate, hence not all the results add up to 100%. Participants could review their answers before submitting them. Website cookies were used to assign a unique user identifier to each client computer. Duplicate database entries having the same user ID were eliminated before analysis. All data collected through the survey were anonymised and not personally identifiable. The online survey’s technical functionality was tested before being published.

Drivers and barriers for the health and social care workforce to promote self-care in individuals living with MM were evaluated by establishing participants’ professional experiences of providing services for individuals with MM. This was explored through questions estimating the proportion of service-users with MM, their most common needs, and the methods used by respondents to promote self-care to manage service-user’s conditions. Several questions on training were also asked, as well as questions exploring perceived gaps in H&CPs education. Most questions were multiple choice, but a variety of formats were used, including statements rated against Likert-style scales. Questions frequently offered the option to add free text clarification.

### Statistical analysis

Quantitative data were collected using an eSurvey questionnaire administered on Qualtrics. Survey responses were summarised using frequencies and percentages. All analyses were performed using SPSS (Statistical Package for Social Sciences) version 28.0.1). The quality of the survey was assessed by completing the Checklist for Reporting Results of Internet E-Surveys (CHERRIES) (26).

### Ethics

The study was reviewed and received a favourable opinion from Imperial College Research Ethics Committee (ICREC # 21IC6770). All data generated or analysed during this study are included in this published article.

### Consent

The first question in the eSurvey was to seek consent. Full information about the study was given and consent was sought from each participant. The right of the participant to refuse to participate without giving reasons was respected. All participants were free to withdraw at any time and without giving reasons. Participants were notified of the publication plans and reminded that all their data was pseudonymised.

### Patient and Public Involvement

No patients were involved. Co-authors Sonia Richardson and John Norton are experienced Public Partners, attached to the NIHR ARC NWL Multimorbidity and Mental Health Theme. They contributed to developing the survey questions, and reviewed the manuscript, critically commenting on the main results and discussion points.

## Results

### Demographic profile of respondents

The total number of respondents was adjusted from 88 to 81, as 7 individuals consented but did not provide responses to the survey question. The survey captured responses from 81 H&CPs in the UK. Most (66.7%, n=48) were female, and the majority (76.4%; n=55) were from a white background. Thirty-seven percent (n=30) were allied health professionals; from whom 13.5% (n=12) were occupational therapists; 20.2% (n=18) were other allied health professionals; 19.1% (n= 17) were from a variety of other occupations which were not disclosed, and 15.7% (n=14) were general practitioners. The remainder were nursing (9%; n=8), social care staff (5.6%; n=5), social prescribers (4.5%; n=4), pharmacists (4.5%; n=4), secondary care consultants (2.2%; n=2), advocacy workers (2.2%; n=2), health coach (1.1%; n=1) health care assistant (1.1%; n=1), and rehabilitation staff (1.1%; n=1).

Not all participants responded to every question hence the total number of responses to each question described in tables 1 and 2 does not always add up to 81. **Table 1**.

**Table 1:** Respondent Characteristics.

|  | N | (%) |
| --- | --- | --- |
| **Age** |  |  |
| 20-29 | 10 | (14.2) |
| 30-39 | 11 | (15.8) |
| 40-49 | 16 | (22.9) |
| 50-59 | 29 | (41.4) |
| 60 & over | 4 | (5.7) |
| **Gender** |  |  |
| Male | 23 | (31.9) |
| Female | 48 | (66.7) |
| Other | 1 | (1.4) |
| **Ethnicity** |  |  |
| White | 55 | (76.4) |
| Black/Black British | 3 | (4.2) |
| Asian/Asian British | 9 | (12.5) |
| Mixed | 1 | (1.4) |
| Other | 1 | (1.4) |
| Prefer not to say | 3 | (4.2) |
| **Occupation** |  |  |
| GP | 14 | (15.7) |
| Nurse | 8 | (9.0) |
| Allied Health Care Professional | 18 | (20.2) |
| Health coach | 1 | (1.1) |
| Social care | 5 | (5.6) |
| Rehabilitation | 1 | (1.1) |
| Social prescriber | 4 | (4.5) |
| Advocacy worker | 2 | (2.2) |
| Health care assistant | 1 | (1.1) |
| Occupational therapy | 12 | (13.5) |
| Pharmacist | 4 | (4.5) |
| Hospital physician | 2 | (2.2) |
| Other | 17 | (19.1) |

**Table 2:**
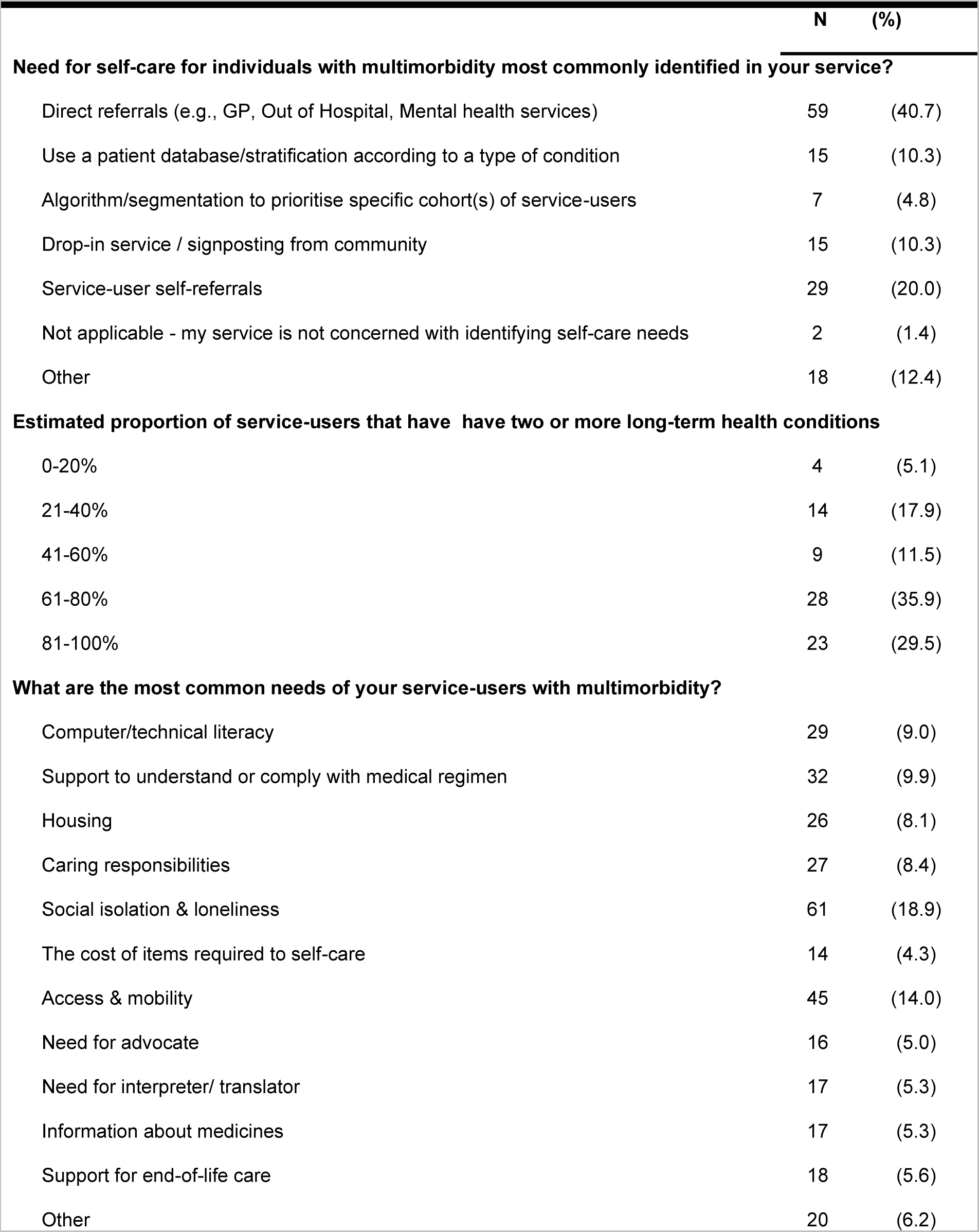
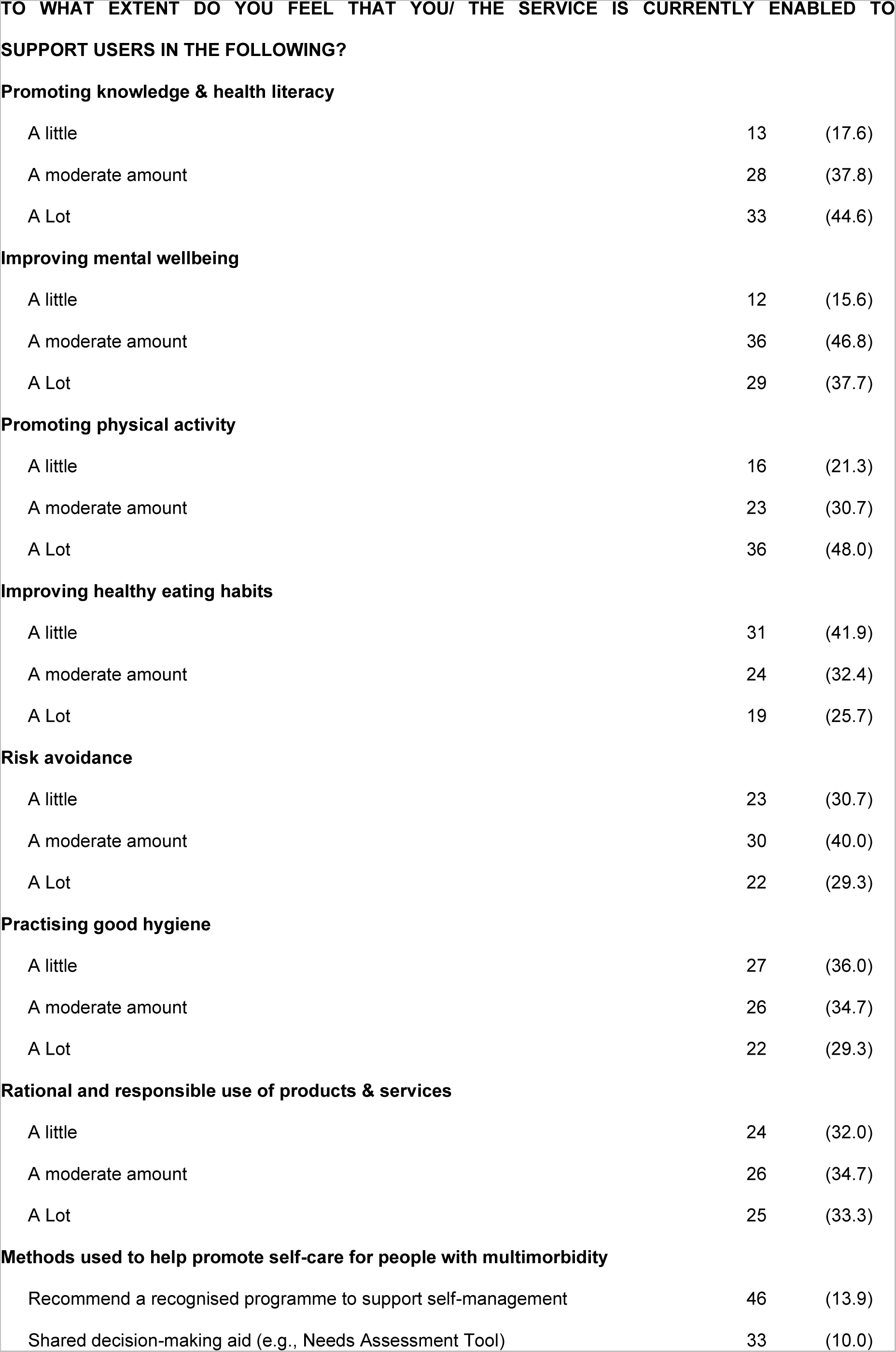
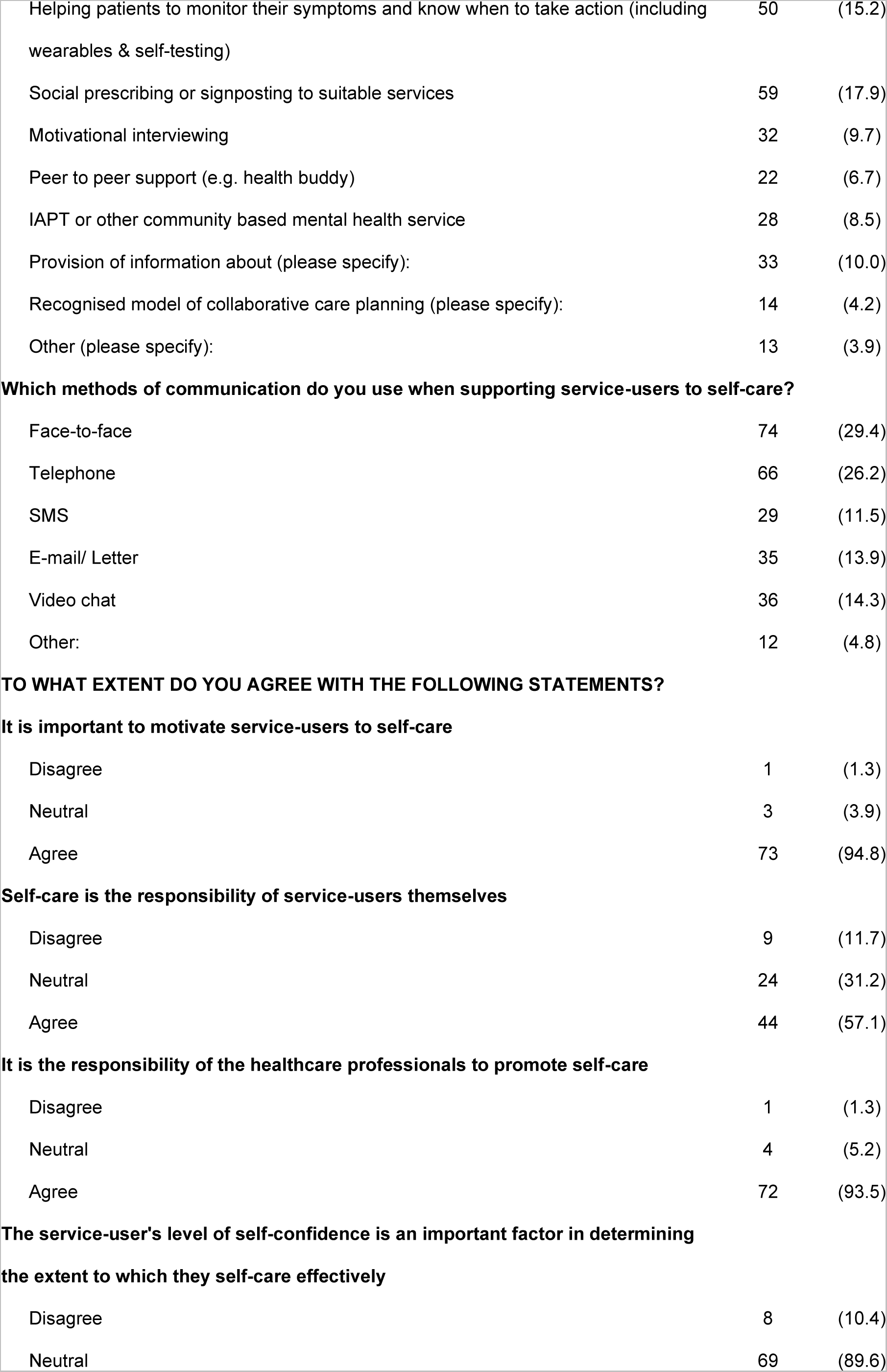
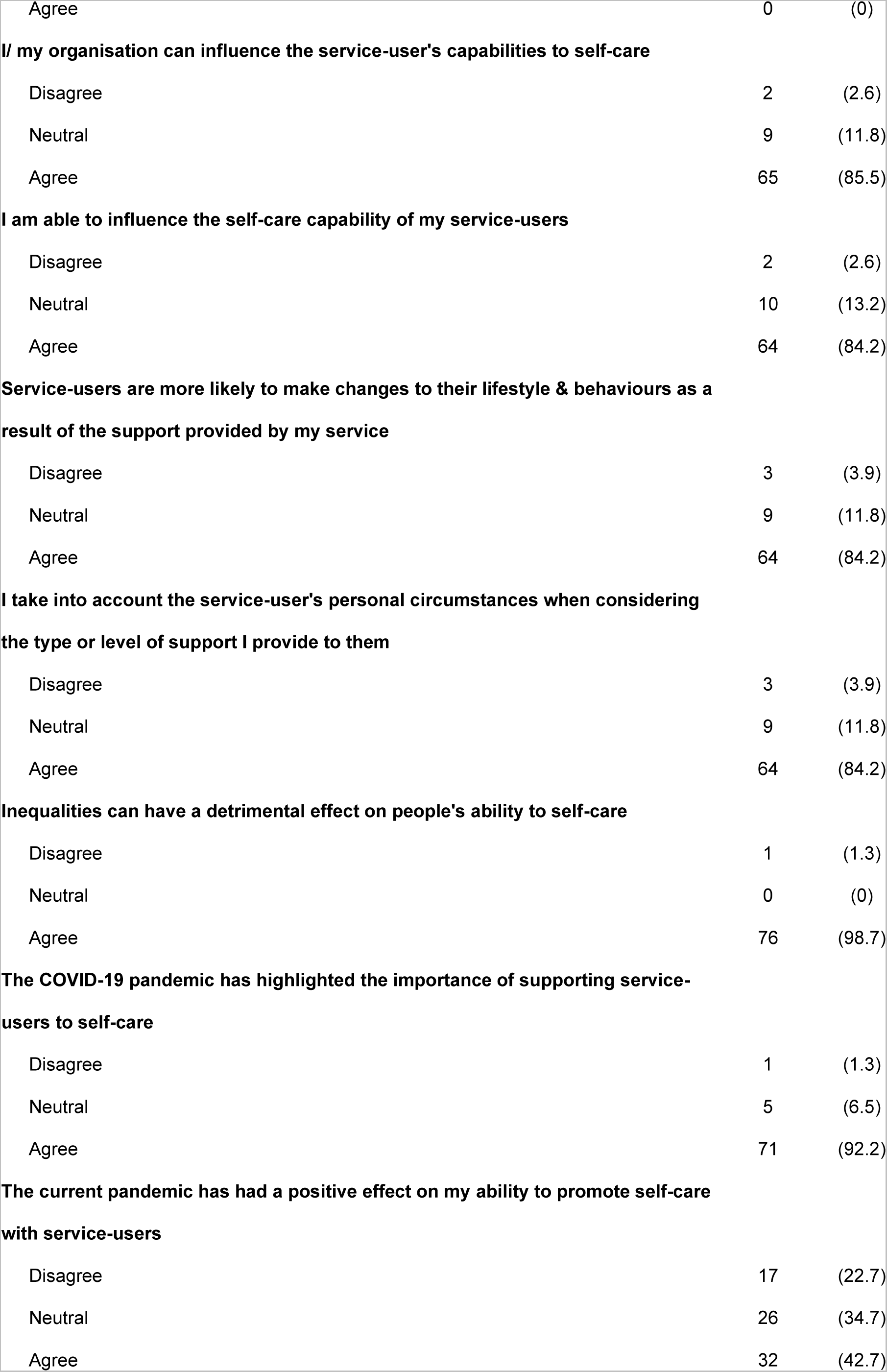
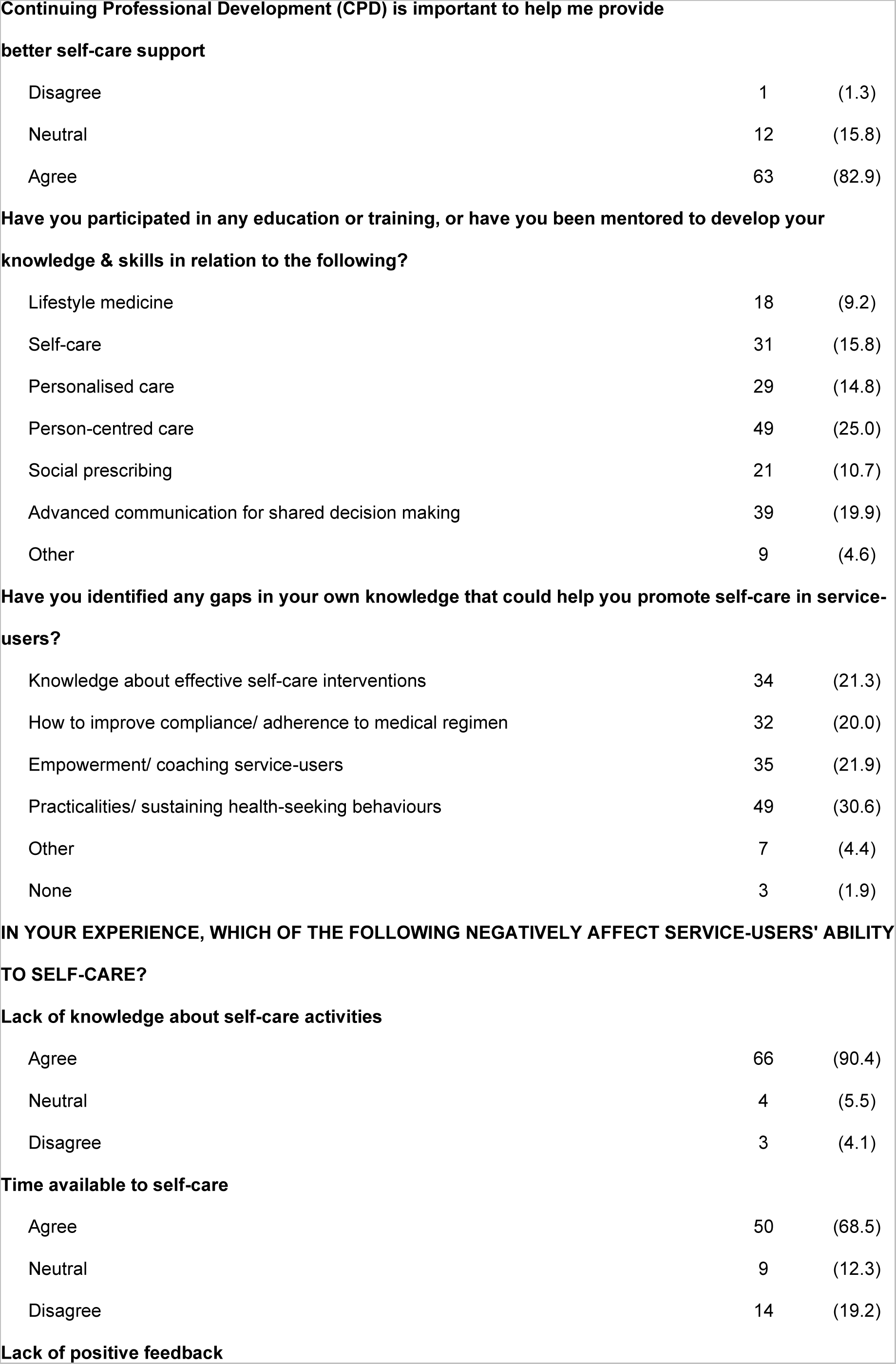
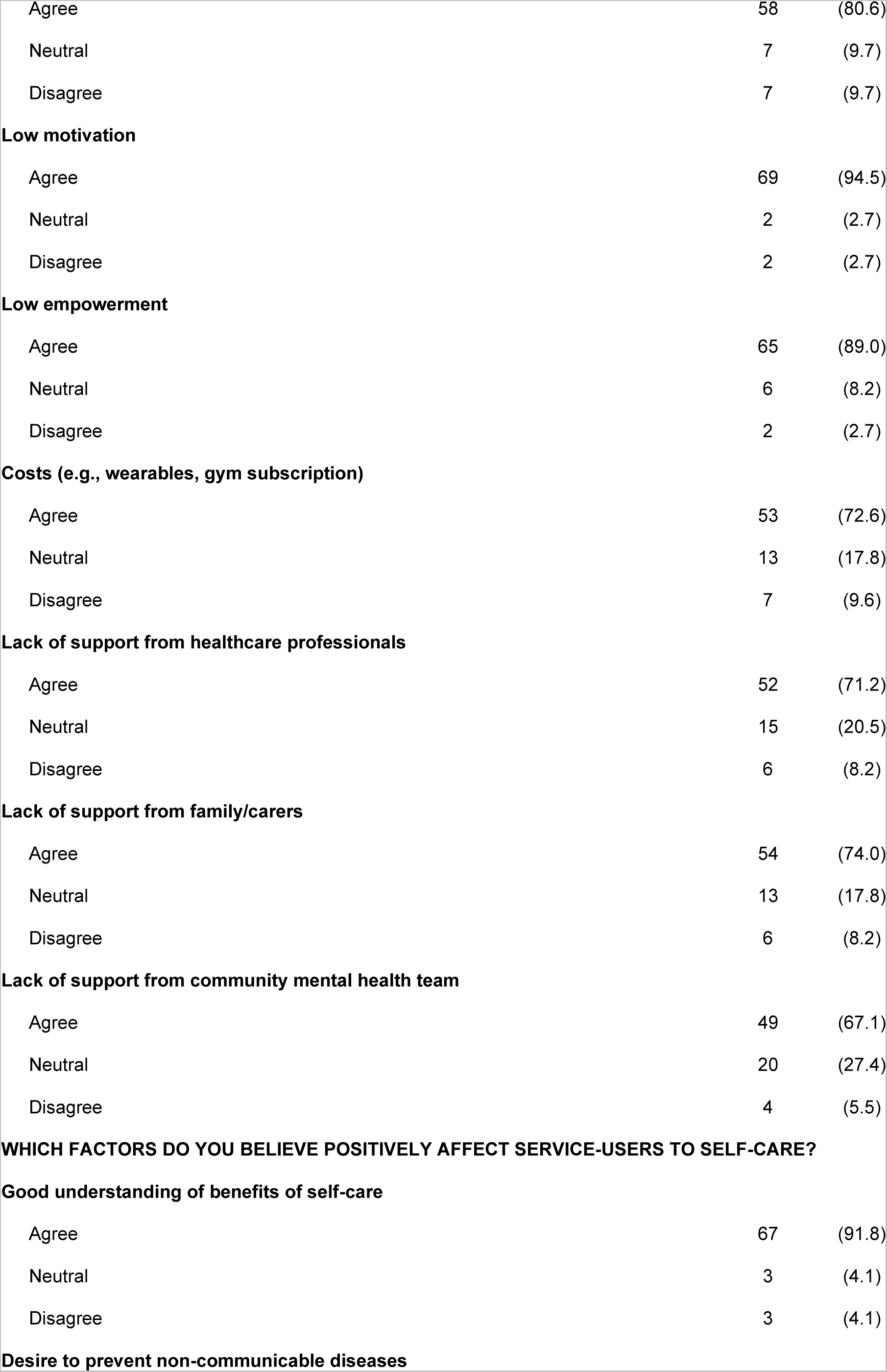
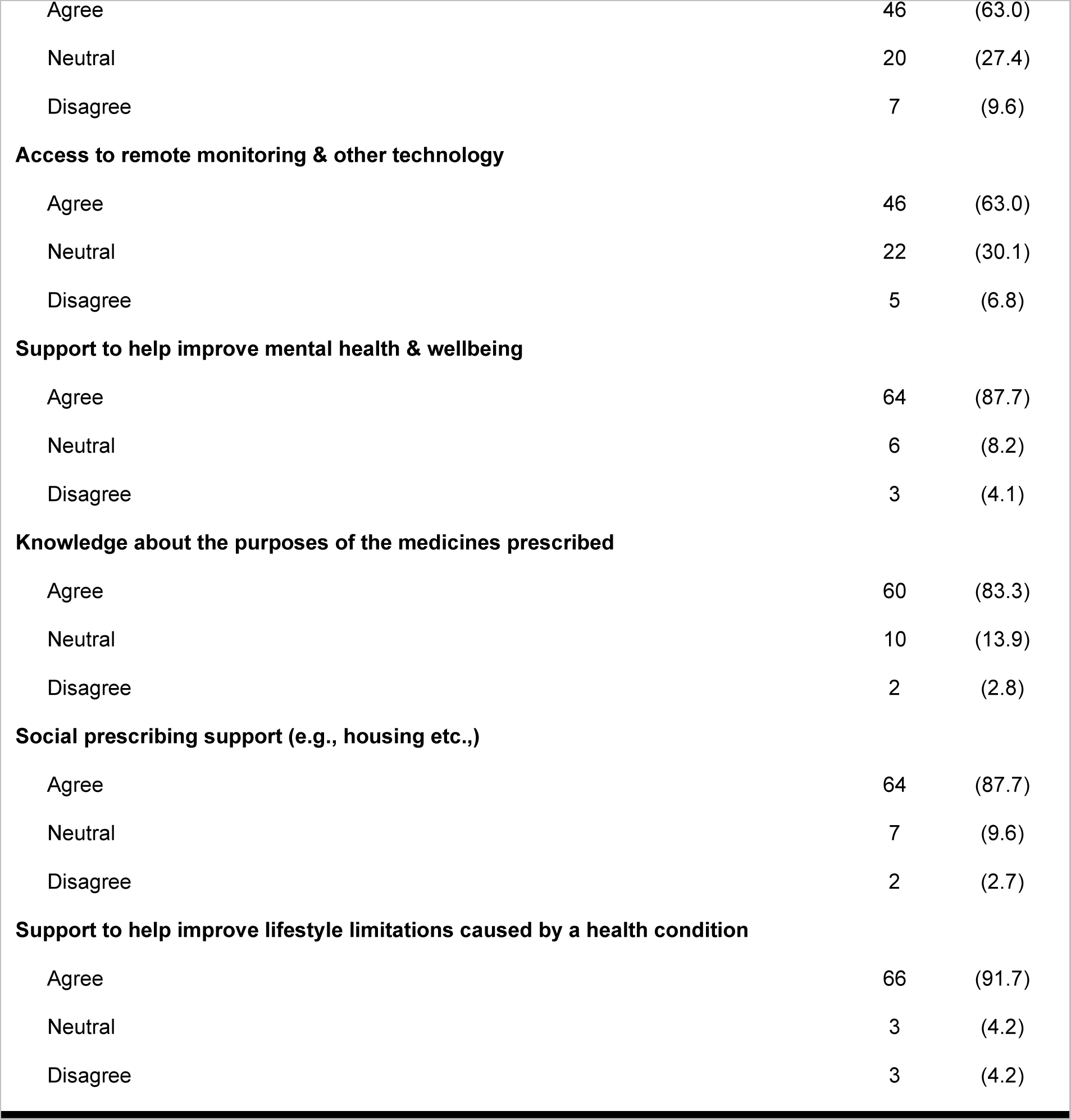
Survey Results.

|  | N | (%) |
| --- | --- | --- |
| **Need for self-care for individuals with multimorbidity most commonly identified in your service?** |  |  |
| Direct referrals (e.g., GP, Out of Hospital, Mental health services) | 59 | (40.7) |
| Use a patient database/stratification according to a type of condition | 15 | (10.3) |
| Algorithm/segmentation to prioritise specific cohort(s) of service-users | 7 | (4.8) |
| Drop-in service / signposting from community | 15 | (10.3) |
| Service-user self-referrals | 29 | (20.0) |
| Not applicable - my service is not concerned with identifying self-care needs | 2 | (1.4) |
| Other | 18 | (12.4) |
| **Estimated proportion of service-users that have have two or more long-term health conditions** |  |  |
| 0-20% | 4 | (5.1) |
| 21-40% | 14 | (17.9) |
| 41-60% | 9 | (11.5) |
| 61-80% | 28 | (35.9) |
| 81-100% | 23 | (29.5) |
| **What are the most common needs of your service-users with multimorbidity?** |  |  |
| Computer/technical literacy | 29 | (9.0) |
| Support to understand or comply with medical regimen | 32 | (9.9) |
| Housing | 26 | (8.1) |
| Caring responsibilities | 27 | (8.4) |
| Social isolation & loneliness | 61 | (18.9) |
| The cost of items required to self-care | 14 | (4.3) |
| Access & mobility | 45 | (14.0) |
| Need for advocate | 16 | (5.0) |
| Need for interpreter/ translator | 17 | (5.3) |
| Information about medicines | 17 | (5.3) |
| Support for end-of-life care | 18 | (5.6) |
| Other | 20 | (6.2) |
| **Promoting knowledge & health literacy** |  |  |
| A little | 13 | (17.6) |
| A moderate amount | 28 | (37.8) |
| A Lot | 33 | (44.6) |
| **Improving mental wellbeing** |  |  |
| A little | 12 | (15.6) |
| A moderate amount | 36 | (46.8) |
| A Lot | 29 | (37.7) |
| **Promoting physical activity** |  |  |
| A little | 16 | (21.3) |
| A moderate amount | 23 | (30.7) |
| A Lot | 36 | (48.0) |
| **Improving healthy eating habits** |  |  |
| A little | 31 | (41.9) |
| A moderate amount | 24 | (32.4) |
| A Lot | 19 | (25.7) |
| **Risk avoidance** |  |  |
| A little | 23 | (30.7) |
| A moderate amount | 30 | (40.0) |
| A Lot | 22 | (29.3) |
| **Practising good hygiene** |  |  |
| A little | 27 | (36.0) |
| A moderate amount | 26 | (34.7) |
| A Lot | 22 | (29.3) |
| **Rational and responsible use of products & services** |  |  |
| A little | 24 | (32.0) |
| A moderate amount | 26 | (34.7) |
| A Lot | 25 | (33.3) |
| **Methods used to help promote self-care for people with multimorbidity** |  |  |
| Recommend a recognised programme to support self-management | 46 | (13.9) |
| Shared decision-making aid (e.g., Needs Assessment Tool) | 33 | (10.0) |
| Helping patients to monitor their symptoms and know when to take action (including wearables & self-testing) | 50 | (15.2) |
| Social prescribing or signposting to suitable services | 59 | (17.9) |
| Motivational interviewing | 32 | (9.7) |
| Peer to peer support (e.g. health buddy) | 22 | (6.7) |
| IAPT or other community based mental health service | 28 | (8.5) |
| Provision of information about (please specify): | 33 | (10.0) |
| Recognised model of collaborative care planning (please specify): | 14 | (4.2) |
| Other (please specify): | 13 | (3.9) |
| **Which methods of communication do you use when supporting service-users to self-care?** |  |  |
| Face-to-face | 74 | (29.4) |
| Telephone | 66 | (26.2) |
| SMS | 29 | (11.5) |
| E-mail/ Letter | 35 | (13.9) |
| Video chat | 36 | (14.3) |
| Other: | 12 | (4.8) |
| **TO WHAT EXTENT DO YOU AGREE WITH THE FOLLOWING STATEMENTS?** |  |  |
| **It is important to motivate service-users to self-care** |  |  |
| Disagree | 1 | (1.3) |
| Neutral | 3 | (3.9) |
| Agree | 73 | (94.8) |
| **Self-care is the responsibility of service-users themselves** |  |  |
| Disagree | 9 | (11.7) |
| Neutral | 24 | (31.2) |
| Agree | 44 | (57.1) |
| **It is the responsibility of the healthcare professionals to promote self-care** |  |  |
| Disagree | 1 | (1.3) |
| Neutral | 4 | (5.2) |
| Agree | 72 | (93.5) |
| **The service-user's level of self-confidence is an important factor in determining the extent to which they self-care effectively** |  |  |
| Disagree | 8 | (10.4) |
| Neutral | 69 | (89.6) |
| Agree | 0 | (0) |
| **I/ my organisation can influence the service-user's capabilities to self-care** |  |  |
| Disagree | 2 | (2.6) |
| Neutral | 9 | (11.8) |
| Agree | 65 | (85.5) |
| **I am able to influence the self-care capability of my service-users** |  |  |
| Disagree | 2 | (2.6) |
| Neutral | 10 | (13.2) |
| Agree | 64 | (84.2) |
| **Service-users are more likely to make changes to their lifestyle & behaviours as a result of the support provided by my service** |  |  |
| Disagree | 3 | (3.9) |
| Neutral | 9 | (11.8) |
| Agree | 64 | (84.2) |
| **I take into account the service-user's personal circumstances when considering the type or level of support I provide to them** |  |  |
| Disagree | 3 | (3.9) |
| Neutral | 9 | (11.8) |
| Agree | 64 | (84.2) |
| **Inequalities can have a detrimental effect on people's ability to self-care** |  |  |
| Disagree | 1 | (1.3) |
| Neutral | 0 | (0) |
| Agree | 76 | (98.7) |
| **The COVID-19 pandemic has highlighted the importance of supporting service-users to self-care** |  |  |
| Disagree | 1 | (1.3) |
| Neutral | 5 | (6.5) |
| Agree | 71 | (92.2) |
| **The current pandemic has had a positive effect on my ability to promote self-care with service-users** |  |  |
| Disagree | 17 | (22.7) |
| Neutral | 26 | (34.7) |
| Agree | 32 | (42.7) |
| **Continuing Professional Development (CPD) is important to help me provide better self-care support** |  |  |
| Disagree | 1 | (1.3) |
| Neutral | 12 | (15.8) |
| Agree | 63 | (82.9) |
| Lifestyle medicine | 18 | (9.2) |
| Self-care | 31 | (15.8) |
| Personalised care | 29 | (14.8) |
| Person-centred care | 49 | (25.0) |
| Social prescribing | 21 | (10.7) |
| Advanced communication for shared decision making | 39 | (19.9) |
| Other | 9 | (4.6) |
| **Have you identified any gaps in your own knowledge that could help you promote self-care in service-users?** |  |  |
| Knowledge about effective self-care interventions | 34 | (21.3) |
| How to improve compliance/ adherence to medical regimen | 32 | (20.0) |
| Empowerment/ coaching service-users | 35 | (21.9) |
| Practicalities/ sustaining health-seeking behaviours | 49 | (30.6) |
| Other | 7 | (4.4) |
| None | 3 | (1.9) |
| **Lack of knowledge about self-care activities** |  |  |
| Agree | 66 | (90.4) |
| Neutral | 4 | (5.5) |
| Disagree | 3 | (4.1) |
| **Time available to self-care** |  |  |
| Agree | 50 | (68.5) |
| Neutral | 9 | (12.3) |
| Disagree | 14 | (19.2) |
| **Lack of positive feedback** |  |  |
| Agree | 58 | (80.6) |
| Neutral | 7 | (9.7) |
| Disagree | 7 | (9.7) |
| **Low motivation** |  |  |
| Agree | 69 | (94.5) |
| Neutral | 2 | (2.7) |
| Disagree | 2 | (2.7) |
| **Low empowerment** |  |  |
| Agree | 65 | (89.0) |
| Neutral | 6 | (8.2) |
| Disagree | 2 | (2.7) |
| **Costs (e.g., wearables, gym subscription)** |  |  |
| Agree | 53 | (72.6) |
| Neutral | 13 | (17.8) |
| Disagree | 7 | (9.6) |
| **Lack of support from healthcare professionals** |  |  |
| Agree | 52 | (71.2) |
| Neutral | 15 | (20.5) |
| Disagree | 6 | (8.2) |
| **Lack of support from family/carers** |  |  |
| Agree | 54 | (74.0) |
| Neutral | 13 | (17.8) |
| Disagree | 6 | (8.2) |
| **Lack of support from community mental health team** |  |  |
| Agree | 49 | (67.1) |
| Neutral | 20 | (27.4) |
| Disagree | 4 | (5.5) |
| **WHICH FACTORS DO YOU BELIEVE POSITIVELY AFFECT SERVICE-USERS TO SELF-CARE?** |  |  |
| **Good understanding of benefits of self-care** |  |  |
| Agree | 67 | (91.8) |
| Neutral | 3 | (4.1) |
| Disagree | 3 | (4.1) |
| **Desire to prevent non-communicable diseases** |  |  |
| Agree | 46 | (63.0) |
| Neutral | 20 | (27.4) |
| Disagree | 7 | (9.6) |
| **Access to remote monitoring & other technology** |  |  |
| Agree | 46 | (63.0) |
| Neutral | 22 | (30.1) |
| Disagree | 5 | (6.8) |
| **Support to help improve mental health & wellbeing** |  |  |
| Agree | 64 | (87.7) |
| Neutral | 6 | (8.2) |
| Disagree | 3 | (4.1) |
| **Knowledge about the purposes of the medicines prescribed** |  |  |
| Agree | 60 | (83.3) |
| Neutral | 10 | (13.9) |
| Disagree | 2 | (2.8) |
| **Social prescribing support (e.g., housing etc.,)** |  |  |
| Agree | 64 | (87.7) |
| Neutral | 7 | (9.6) |
| Disagree | 2 | (2.7) |
| **Support to help improve lifestyle limitations caused by a health condition** |  |  |
| Agree | 66 | (91.7) |
| Neutral | 3 | (4.2) |
| Disagree | 3 | (4.2) |

### Service-user identification and their most common needs

More than half (65.4%; n=51) of H&CPs estimated that their patients had multimorbidity (MM), defined as two or more long-term health conditions. Most respondents (65.4%; n=70) provided services to identify and support self-care for people living with MM. Respondents indicated that almost half of service-users (40.7%; n=59) were referred directly to the service, whilst 20% (n=29) engaged with the provider following self-referral.

The needs that were most associated with service-users with MM were related to loneliness and social isolation (18.9%; n=61), mobility and access issues (14%; n=45), and support to understand or comply with a medical regimen (9.9%; n=32).

### Factors affecting service-users’ ability to self-care

Factors perceived to positively affect service-users’ ability to self-care included: a good understanding of the benefits of self-care (91.8%; n=67), receiving support to help improve lifestyle limitations caused by physical/mental health conditions (91.7%; n=66), receiving support to improve mental health and wellbeing (87.7%; n=64) and receiving support via services such as social prescribing and to address unmet social needs, for example, housing (87.7%; n=64).

Factors perceived to impact service-users’ ability most negatively to self-care included low motivation (94.5% n=69), lack of knowledge about self-care activities (90.4% n=66), and low empowerment (89%; n=65). Additionally, (80.6% n=58) indicated that a lack of positive feedback had a negative impact on service user’s ability self-care. Many respondents highlighted a lack of support from family/carers (74%; n=54), healthcare professionals (72.6%; n=53), the costs associated with self-care such as gym membership or wearable aid (67.1%; n=49), a lack of support from a community health team (66.8%; n=50) or the time needed to self-care (68.5%; n=50) all had a negative impact.

### Services and strategies used to support self-care

Respondents signposted service-users to other services, including social prescribing (17.9%; n=59), information services, including digital health technologies (e.g., wearables) and decision support tools (e.g., self-testing) to help individuals monitor their own symptoms (15.2%; n=50) and other recognised programmes to support self-management (13.9%; n=46).

Over half of service providers reported that they mostly communicated with service-users either face to face (29.4%; n=74) and/or by telephone (26.2%; n=66), with fewer communicating by video chat (14.9% n=36), email or letter (13.9% n=35) SMS, (11.5% n=29), other means of communication (4.8% n=12).

### Extent that health and care professionals could support people with multimorbidity to adopt activities across the Seven Pillars of Self-Care

Most respondents felt that they and/or their organisations were enabled to support service-users “a lot” (44.6%; n=33) or by a moderate amount (37.8%; n=28) in improving health literacy levels. Respondents also felt that health and care staff were able to improve service users’ mental wellbeing to varying degrees (37.7%; n=29 by a lot and 46.8%; n=36 by a moderate amount), promoting physical activity (48%; n=36 by a lot, and 30.7%; n=23 by a moderate amount), improving healthy eating habits (25.7%; n=19 by a lot, and 32.4%; n=24 a moderate amount), avoiding risks (29.3%; n=22by a lot, and 40%; n=30 to a moderate extent), improving hygiene practices (29.3%; n=22 by a lot, and 34.7%; n=26 a moderate amount), and promoting the rational use of products and services 33.3%; n=25 a lot, 34.7% (n=26) a moderate amount.

### Perceptions of responsibility and influence on the behaviour of service-users

Most respondents agreed with the following statements: it is important to motivate service-users to self-care (94.8%; n=73); it is the responsibility of H&CPs to promote self-care (93.5%; n=72); they (84,0%; n=64) and their organisation (85.5%; n=65) are able to positively influence the self-care capability of service-users; service-users are more likely to make changes to their lifestyle and behaviours as a result of the support provided (84.2% n=64); H&CPs take account of service-user’s personal circumstances when considering the type or level of support offered (84.2%; n=64). Most respondents indicated that inequalities have a detrimental effect on people’s ability to self-care (98.7%; n=76).

### Impact of the COVID-19 pandemic

Almost all respondents (92.2%; n=71) felt that the pandemic had highlighted the importance of supporting service-users to self-care, whereas nearly half (42.7%; n=32) felt that the pandemic had a positive effect on their ability to promote self-care in service-users.

### The importance placed on continuing professional development

The majority of respondents (82.9%; n=63) indicated that continuing professional development was a key factor in helping them to provide better support to service-users to enable self-care, but only a quarter of respondents (25.0%; n=49) had participated in education and/or training to support person-centred care, or shared decision making (19.9%; n=39). Only a small proportion of respondents had participated in training or received mentoring relevant to self-care (15.7% n=31), personalised care (14.5%; n=29), social prescribing (10.7%; n=21) or lifestyle medicine (9.2%; n=18).

Many respondents identified gaps in their knowledge, especially in relation to improving their understanding and skills to support service-users to sustain health seeking behaviour (30.6%; n=49), or coaching service-users to self-care (21.9%; n=35). When ranking areas of knowledge for improvement, respondents said that knowledge about self-care interventions, empowerment and coaching service-users, and the practicalities of sustaining health seeking behaviours were the most important to them.

## Discussion

### Summary of principal findings

The majority of health and care professionals (H&CPs) identified mobility challenges, loneliness, social isolation, and service access as key obstacles to self-care in service-users. The service-users’ knowledge and understanding is a critical factor for successfully adopting self-care practices, and we reported on the widespread consensus that also emerged on how societal inequalities adversely affect self-care capabilities. Positive influences on self-care efforts and outcomes included support from family, carers, or professional services. In contrast, barriers like low motivation, insufficient knowledge, lack of encouragement, and inadequate support, alongside the costs and time involved, hinder the routine uptake of self-care behaviours outside clinical environments.

H&CPs identified their strength in employing person-centered methods to boost health literacy, mental health, hygiene, physical activity, and the rational use of health products and services among service-users. Yet, only a few believed their interactions significantly influenced long-term healthy eating habits in service-users.

Continued professional development was deemed crucial, and whilst many H&CPs have received training on supporting self-care, gaps remain, particularly in strategies for practical engagement and maintaining health-seeking behaviours, such as medication adherence and the need for more evidence-based and coaching-style methods to enhance self-care skills.

### Interpretation and comparison with literature

The COVID-19 pandemic underscored the importance of self-care, with about half the participants noting that pandemic-induced care model changes improved their support for self-care. Recent findings suggest the pandemic altered H&CPs’ attitudes toward self-care, advocating for its inclusion in healthcare education to sustain these attitude shifts and boost individual empowerment in self-care. This finding is consistent with the results of another study that characterised experience of the pandemic and found that it had changed healthcare professional’s attitudes to self-care (27).

Earlier studies on self-care provide useful learning and detail positive outcomes from interventions to improve self-care relating to long-term conditions such as type 2 diabetes (28) and specific co-morbidities such as cardiovascular disease and stroke (29), depression, diabetes, and heart disease (7). There are fewer studies that investigated the impact of supported self-care on service-users with more diverse multimorbidity. Earlier studies tend to focus on patient outcomes, rather than knowledge, attitudes, and perceptions of H&CPs. Thus, in the study reported in this paper, H&CPs reported the potential to positively influence the lifestyle and behaviour of service users in relation to certain behaviours, including exercise and good hygiene, there was less certaintly reported regarding the extent that lifestyle advice could help improve healthy eating habits. This contrasts with studies of supported self-management that reported significant positive outcomes for people with multimorbidity in relation to diet (8). This may be attributed to the small number of respondents who were dieticians.

Social isolation, loneliness, a perceived lack of support to tackle lifestyle factors such as low motivation, poor knowledge about self-care activities, time constraints and associated costs could negatively impact our ability to self-care. H&CPs indicated the potential detrimental impact of inequalities on service-users with multimorbidity, and this likely underpins additional challenges for both the person with multimorbidity and the service provider. Our findings are consistent with the results of a recent study that considered the impact of inequalities on the outcomes of self-care interventions in eight primary care locations among individuals with multimorbidity in Canada (30). Of the six lifestyle factors under consideration, the likelihood of improvement was no longer significant for people on lower incomes and with lower educational status in the domains of emotional wellbeing, self-monitoring and insight (30).

As other studies show, knowledge, training and preparedness on the part of H&CPs are vital to supporting self-care (13). This includes training of primary care staff on co-designing supported self-care plans which resulted in enhanced team working and the promotion of necessary behaviour change among service-users (31). Some studies identified the need for more H&CP disease-specific training alongside training to support self-care (10,32). This highlights the need to explore more opportunities to promote truly integrated care approaches by developing training and care resources with greater involvement from multidisciplinary teams and patients who live with multimorbidity (33,34). This approach can help embed learning and best practice so that patient experience and services can be improved to account for what matters most to service users (33,34).

### Implications for clinicians and policy makers

Supported self-care is part of the NHS commitment to make personalised care business as usual across the health and care system (18,25). Supported self-care interventions are most successful if delivered as part of integrated care interventions for people living with long-term health conditions (35). This highlights the need for a shift in the relationship between H&CPs and service-users, in which the health conditions and needs of the whole person are considered together (36), and in which that person’s own skills, strengths and attributes are considered (37). Guidelines on supporting self-care are designed to support service providers to care for people with long-term health conditions and start from the premise that people want to have choice and control over the way their care is planned and delivered, based on what matters to them and their individual strengths, needs and preferences (18).

H&CPs reported that continuing professional development was important and some had experience of relevant training. However, the number that had completed training was small, and gaps in knowledge about how to improve practical interactions to sustain health-seeking behaviours were reported. This suggests that H&CPs require more choice of training or experience to help them to promote the uptake and sustained adoption of self-care among service-users. In the UK, NHS Learn and the Personalised Care Institute aim to address this gap by helping H&CPs to develop the knowledge and skills to support the delivery of the comprehensive model for personalised care which as a domain has a direct bearing on the self-care (38).

Additional training, learning on the job and mentoring may improve the knowledge and skills base of H&CPs. This could be linked to the development of clearer clinical guidelines (39,40), teamworking and developing a broader consensus on the key elements of self-care likely to bring positive changes (10,13,31,32,41) and could be achieved by bringing resources from services across the public sector to shape and reduce the negative impacts of the social determinants of health (42).

### Strengths and limitations

To our knowledge, this is the first study in the UK that sought to investigate the knowledge, attitudes and perceptions of health and care professionals in the UK concerning extant barriers and drivers to promote self-care among service-users with multimorbidity.

A key strength of our study is that it provided insights from a variety of service providers about what barriers continue to be challenging to individuals with multimorbidity and to the health and care professionals themselves. To our knowledge, only one other study has sought to investigate the prevailing attitude of H&CPs on self-care following the advent of the COVID-19 pandemic (27). This considered a general population of service users, while we specifically focused on a cohort of service users who live with multimorbidity. The main limitation of our study was that it only collected data from 81 respondents and for this reason, we cannot assume that the cross-sample is representative of the health and care workforce. Due to the nature of the online survey obtaining a response rate was not possible. However, the results do demonstrate that the study attracted a broad representation of roles from a range of health and social care professionals, offering a comprehensive view of the systemic challenges and facilitators in promoting self-care in the context of multimorbidity.

Understanding the needs of individuals who have multimorbidity and the needs of H&CPs who care for them is important. Our study asked H&CPs about the extent to which they were able to meet service-user’s needs, the type of training they had completed and outstanding gaps in their training and experience. In addition, it elicited their perceptions about the role of service-users, and the factors that impact on service-users ability to self-care. This is important in the current policy and service provision context in which the agency of individuals with multimorbidity must be taken account of in supporting the development and use of person-centred/personalised care models (19,26). The study highlighted gaps in knowledge among service providers, which, if addressed, would improve their ability to support people with multimorbidity and would be likely to improve self-care and potentially health outcomes.

### Outstanding questions/Implications for research

More research is required to better understand the types of training or other methods of skill and knowledge acquisition by H&CPs, which could improve the support available to people living with multimorbidity. Future research could usefully focus on what might equip H&CPs to become more knowledgeable and confident in choosing strategies and interventions designed to improve motivation and meet service-users’ needs.

The strategies and techniques most frequently used by respondents to our survey were social prescribing, helping service-users monitor their symptoms and know when to act, with a majority indicating the importance of service-user motivation in behaviour change. Social prescribing was designed to tackle challenges and put person-centred care centre stage. A recent study suggested that there has been limited exploration of the impact of social prescribing (43). A recent systematic literature review reported inconclusive evidence for social prescribing as an effective approach to improving health outcomes for adults experiencing multimorbidity and social deprivation in primary and community care settings (44). Further research could explore the drivers and barriers to optimal outcomes desired from social prescribing by H&CPs and service-users.

Additionally, more research into the most optimal use of communication channels may be useful. Our study suggested that communication with service-users was mostly face-to-face and by telephone, but respondents also reported the use of new communication tools e.g., video chat. New research could explore the extent to which H&CPs can influence service-user’s ability to self-care if delivered remotely, and support remain person-centred and pro-active care.

## Conclusions

Multimorbidity is increasingly becoming a global concern, primarily due to the ageing population and lifestyle changes. Knowledge and understanding on the part of service-users with multimorbidity is key to successful engagement with and uptake of evidence-based self-management and self-care interventions. Policymakers should therefore focus on ensuring that appropriate information, training, mentoring and support is available to health and care professionals and patients. The pandemic appears to have shed light on the important role that self-care could play in patients with multimorbidity, and the ensuing changes in the models of care observed may have resulted in improvements in health and care professionals’ ability to support service-users in their personal self-management and self-care journey. More research is needed to understand changes resulting from the pandemic and their impact, and how these lessons can be applied to promote the sustained adoption of health-seeking self-care behaviours to tackle the rise of non-communicable diseases and multimorbidity.

## Author Contributors

All authors provided substantial contributions to the conception design, acquisition and interpretation of study data and approved the last version of the paper. AEO took the lead in planning the study with support from co-authors. Susan Barber took the lead in drafting the manuscript with critical review and input from Benedict Hayhoe. Sonia Richardson and John Norton contributed to the survey questions and reviewed the first and final drafts of the manuscript for publication. Manisha Karki carried out the data analysis with support from Austen El Osta, Benedict Hayhoe and Susan Barber. Austen El Osta is the guarantor.

## Data Availability

All relevant data are within the manuscript and its Supporting Information files.

## Acknowledgements

The authors thank Iman Webber and Aos Alaa for supporting development of the survey, and for disseminating study information to survey respondents.

## Patient and Public Involvement

No patient was involved in the study. Co-authors Sonia Richardson and John Norton (deceased March 2023) are lay researchers who have direct and indirect experience of multimorbidity and who helped to construct and review the survey questions used in this study.

## Data sharing statement

The data that support the findings of this study are available from the corresponding author, Susan Barber, upon request.

